# Cardiovascular genetic counseling is associated with improved patient-reported outcomes across clinical indications and settings

**DOI:** 10.1101/2025.09.29.25336936

**Authors:** Brittney Murray, Catherine Gordon, Susan Christian, Tara Dzwiniel, Crystal Tichnell, Emily Brown, Gretchen MacCarrick, Rebecca McClellan, Chloe Reuter, Hannah Ison, Jennefer Carter, Tia Moscarello, Ryan Murtha, Mitchel Pariani, Julia Platt, Christina Rigelsky, Diane Clements, Paul Crawford, Joseph Liu, Elizabeth Jordan, Robyn J. Hylind, Eugene K. Wong, Sarah Wang, Zoe Lindsay-Mills, Megan Betts, Eric Tricou, Amy C. Sturm, Adam H. Buchanan, Sara Fitzgerald-Butt, Benjamin M. Helm, Laura Yeates, Grad Dip Gen Couns, Colleen Caleshu, Jodie Ingles, Grad Dip Gen Couns, Lisa Yanek, Cynthia A. James

## Abstract

**Background:** Contemporary guidelines recommend genetic counseling for potentially inherited cardiomyopathies, channelopathies, aortopathies, and dyslipidemias, but few, small, studies characterize cardiovascular genetic counseling (CVGC) outcomes impeding efforts to improve CVGC service delivery. We aimed to 1) quantify psychosocial CVGC outcomes, 2) identify clinical, demographic, and service delivery characteristics associated with extent of benefit from CVGC, and 3) determine how outcomes are associated with patient-reported quality of the genetic counselor (GC):patient alliance.

**Methods:** 890 adults attending a first outpatient CVGC appointment at eight United States and Canadian medical centers completed questionnaires assessing empowerment (Genetic Counseling Outcome Scale [GCOS-24]), worry (modified Cancer Worry Scale [CWS]), cardiac anxiety (Cardiac Anxiety Questionnaire [CAQ]), and GC:patient alliance (Working Alliance Inventory–patient [WAI-SR]) prior to and up to 4 weeks post-CVGC, but before results of genetic tests ordered were returned to decouple CVGC and genetic test result impact.

**Results:** Following CVGC, empowerment increased (GCOS-24 change: +5.94±11.6, p<0.001), worry decreased (CWS change: −0.40±3.55, p=0.002), and cardiac fear decreased (CAQ fear subscale change: −0.034±0.46, p=0.033) with no change in whole-scale CAQ (0.013±0.34, p=0.29). In linear regression, worry decreased most in patients with less formal education (p=0.0021), with an arrhythmia indication (p=0.013), and who were at-risk family members (p=0.016) while strength of the GC:patient alliance had no impact. In contrast, the GC:patient alliance was strongly positively associated with change in empowerment (p<0.001).

**Conclusions:** This observational, multicenter, multiprovider study provides foundational evidence that CVGC is associated with increased empowerment and decreased worry in adults attending outpatient CVGC and suggests opportunities for optimizing CVGC services.

## INTRODUCTION

Genetic counseling and genetic testing are standard of care for families with suspected inherited heart disease (IHD). Contemporary guidelines recommend that genetic counselors (GCs) be included as members of the multidisciplinary care team for these families and that genetic counseling accompany genetic testing^1–3^. Genetic testing is now recommended to inform diagnosis and personalized management for a growing number of cardiovascular indications: cardiomyopathies, arrhythmias, dyslipidemias, and aortopathies^1,4–6^. As in other genetic counseling sub-specialties, this precision medicine era has brought a rapid expansion in the number of patients seeking cardiovascular genetic counseling (CVGC), with providers exploring a variety of alternative genetic counseling service models to accommodate this volume^7–9^. This has been stymied in part by critical gaps in the published CVGC evidence base.

While cardiovascular GCs have long been valued for their expertise in ordering and interpreting genetic tests and attending to the psychosocial needs of families, there has historically been limited sub-specialty-specific evidence to support CVGC practice^4,10^. Most genetic counseling outcomes data has been derived from oncology settings^11^ with few, primarily single-center studies quantifying CVGC outcomes^12,13^ and fewer still that distinguish between genetic counseling and genetic testing outcomes. Emerging data suggest that CVGC improves multifaceted, but especially psychosocial, patient outcomes for IHD families ^12–14^.

However, these studies included limited cardiovascular clinical indications and few practicing GCs, so the generalizability of these findings is unknown. It remains poorly understood which patient, clinical, and service delivery characteristics are associated with the greatest, and least, benefit from CVGC, and vitally, which outcomes are associated with quality of the GC:patient relationship (i.e., the GC:patient alliance)^15^. Addressing these gaps is needed both to plan and evaluate how to serve IHD patients and families in the genomics era.

Quantifying the impact of genetic counseling, separate from genetic test impact, remains challenging. The genetic counseling community has embraced the construct of “empowerment” as a genetic counseling outcome^16,17^. Empowerment is a construct first published in relation to genetics services by McAllister et al. who defines it as ‘a set of beliefs that enable a person from a family affected by a genetic condition to feel that they have some control over and hope for the future^17^.’ In a preliminary study, we showed that CVGC, especially with a strong GC:patient alliance, was associated with increased empowerment^13^. These findings are particularly important because the literature suggests that many of the tenets of genetic counseling and empowerment are associated with patient psychosocial adjustment, medical adherence, and favorable health behaviors, which are highly relevant in cardiac conditions where adherence to medical recommendations can mean avoidance of sudden cardiac death ^18–21^. In the previous study, it was shown that extent of change in empowerment was independent from whether a genetic test was ordered or already received prior to CVGC, providing additional evidence for the improved outcomes specific to the interaction with the GC, not just the genetic test itself ^13^. Nonetheless these findings reflected a single setting, GC, and clinical indication.

It has been consistently shown that families with IHD have elevated, often clinically significant levels of psychological distress including high anxiety^22,23^. In the oncology genetic counseling literature, there is evidence that genetic counseling is associated with reduced anxiety and worry^24,25^. Testing whether CVGC results in a similar reduction across multiple centers, clinicians and a variety of IHD types is informative given that high anxiety in IHD patients contributes to negative health outcomes^26^

Thus, we initiated the Multicenter Cardiovascular Genetic Counseling Outcomes Study (MCVGC-GO) to 1) measure the impact of CVGC on empowerment, worry, and anxiety, 2) identify which clinical, demographic, and CVGC service delivery characteristics were associated with extent of benefit from CVGC, and 3) determine how these CVGC outcomes are associated with the strength of the GC:patient alliance.

## METHODS

### Summary

The MCVGC-GO was a prospective, observational, pre-post study of psychosocial outcomes of CVGC performed by board certified/eligible GCs at eight centers in the United States and Canada with established CVGC services. Adults attending a first outpatient CVGC visit completed questionnaires before and after CVGC with post-counseling questionnaires completed prior to return of any genetic test results ordered during the visit. This approach allows CVGC and genetic test outcomes to be decoupled. Local IRB/Ethics approval was obtained by each MCVGC-GO center. Because of the sensitive nature of the data collected and due to restrictions on data sharing the full dataset will not be made available. Requests to access limited data elements from qualified researchers trained in human subject confidentiality protocols may be sent to the corresponding author.

### Participants

MCVGC-GO centers included Johns Hopkins University, Stanford University, the Cleveland Clinic, Ohio State University, Massachusetts General Hospital, Boston Children’s Hospital, and Geisinger – all in the United States - and the University of Alberta in Canada. Participating centers were required to employ at least one American Board of Genetic Counseling (ABGC) Board Certified or Board-Eligible GC regularly providing outpatient CVGC.

Adults scheduled to attend a first outpatient CVGC visit for a cardiovascular indication at a MCVGC-GO center were eligible to participate. Patients with clinical diagnoses indicative of (possible) IHD and individuals at-risk based on family history and/or familial genetic test results were eligible to participate. Likewise, patients scheduled for in-person or video-based telemedicine visits were eligible. Individuals who had prior genetic testing were also eligible so long as there was no history of CVGC by a GC. Parents attending CVGC primarily for a minor child’s indication were eligible. Individuals with prior CVGC, including inpatient CVGC, or who were unable to speak or read English were ineligible.

### Study outcomes and measures

#### CVGC Outcomes

There is no universally recognized metric or single validated measure for quantifying outcomes of CVGC. We selected outcomes based on 1) findings of prior CVGC outcomes studies, 2) the Reciprocal Engagement Model of genetic counseling, and 3) the literature on individual and familial adaptation to IHD^12,13,22,23,27–29^.

Primary MCVGC-GO outcomes were extent of change in empowerment as measured by the Genetic Counseling Outcome Scale [GCOS-24]) and change in cardiac worry measured via a modified Cancer Worry Scale [CWS]). The GCOS-24 is a 24-item scale with 7 response options per item. Scores range from 24-168, with higher scores indicating higher empowerment. This scale, which was developed and validated in British general genetics patients, evaluates dimensions of perceived personal control, emotional regulation, and hope. It has good internal consistency (α=0.87) and test-retest reliability (r=0.86) and has been used across genetic counseling specialties including CVGC^12,13,17^. The CWS is an 8-item measure with total scores ranging from 8-32, with a higher total score indicating greater worry and/or greater effect on mood and daily functioning because of worry. It has been widely used in studies of genetic testing and counseling in oncology^30^. There is no equivalent scale appropriate for use in a broad cardiovascular or IHD population. Thus, the CWS was modified substituting “heart disease” for “cancer.” We also measured cardiac anxiety as a secondary outcome via the Cardiac Anxiety Questionnaire (CAQ), an 18-item measure with three subscales: avoidance, attention, and fear with good whole scale (α=0.83) and subscale (α=0.83, 0.82, 0.69) reliability^31^. Scores range from 0-4 with higher scores indicating greater heart focused anxiety^32,33^. This validated measure was included as it has been previously utilized in studies of the psychosocial well-being of families with IHD and as a CVGC outcome^13,33,34^. However, there are recognized limitations of using this scale in patients with some genetic cardiovascular indications as avoidance behaviors due to compliance with medical recommendations cannot be easily separated from avoidance due to anxiety. Thus, we selected the modified CWS as the primary outcome.

#### Clinical, demographic, and service delivery characteristics

Demographics were self-reported on the pre-visit questionnaire and included: age, gender, education, and race. Clinical indication was based on report of the treating GC and included type of cardiac condition (dyslipidemia, cardiomyopathy, arrhythmia, aortopathy), whether the patient had a personal history of heart disease, and whether there was a recent sudden death in the family precipitating the visit. The “aortopathy” designation included a range of both aortic and arterial aneurysm or dissection referrals. “Arrhythmia” included atrial and ventricular arrhythmia or sudden cardiac death / arrest with no known cardiomyopathy as well as diagnosed (familial) channelopathies or arrhythmia syndromes. CVGC visit variables included aspects of service delivery and counseling model including: (1) whether the visit was in-person or a video visit, (2) whether the visit was conducted solely by a GC or combined with a physician visit, (3) whether a GC trainee conducted part of the counseling during the session, (4) whether genetic testing was ordered during the visit, and (5) whether the patient came to the visit with a genetic test result. A CVGC visit could include pre- and post-test counseling if the patient presented with a genetic test result and additional genetic testing was then ordered.

#### Genetic counselor:patient alliance

The quality of the GC:patient alliance was measured using the Working Alliance Inventory–client short form (WAI-SR), a 12-item Likert scale^33,35^. This scale has three components (tasks, goals, bonds) that together help define the quality of a working alliance^36^. Scores range from 12-60 with higher scores indicating a stronger alliance. Minor revisions were made replacing “therapist” with “genetic counselor” to adapt this measure for genetic counseling as done previously^13^.

Questions developed by Ison et al. regarding CVGC patients’ awareness of recommendations for personal cardiovascular care and screening of family members were also included in the study questionnaires but are not reported herein^12^. Study questionnaires are available in the **Supplemental Material**.

### Data collection

MCVGC-GO enrollment began 4/15/2020 and ended 8/31/2022, with each center collecting data for 12-18 months. All data were collected after the COVID-19 pandemic had begun and all centers offered video-based telemedicine for CVGC during the enrollment period. Recruitment, questionnaire administration, and collection of CVGC session data were performed locally. At each center, adults scheduled for an eligible CVGC appointment were invited to participate. Two weeks prior to the scheduled CVGC appointment, participants were provided with links to an online pre-visit questionnaire. Reminders were sent one week and 2-3 days prior to the CVGC appointment. Pre-visit questionnaires could be completed up to the day of the scheduled appointment. Patients who completed the pre-visit questionnaire were sent links to the post-visit questionnaire within 4 days after their CVGC appointment. Up to two reminders to complete this questionnaire were sent. While participants were able to complete the post-visit questionnaire up to 4 weeks post-CVGC, participants were no longer eligible to complete post-visit questionnaires once results of genetic testing ordered in conjunction with the CVGC appointment were disclosed. Participants who completed both the pre- and post-CVGC questionnaires were entered into a lottery for $50 gift card incentives at eligible centers. Deidentified data was shared with the MCVGC-GO coordinating center (Johns Hopkins) for analysis.

### Statistical analyses

All scales were scored according to published standard procedures. If any item in a scale was missing, that scale/subscale was not scored. Demographic, clinical, and CVGC characteristics were analyzed using descriptive and frequency statistics. Scale and subscale scores were similarly summarized. For the primary and secondary outcomes, change scores were calculated by comparing the net change between the pre-CVGC scale score and the post-CVGC score. The normality of all continuous variables, including change scores, was tested by the Kolmogorov–Smirnov test. The extent of change in scale and subscale scores pre-post CVGC were assessed via paired t-tests. Bivariate associations between continuous variables and categorical variables were tested using t-tests or ANOVA. To identify demographic, clinical, and CVGC characteristics independently associated with study outcomes, all potential predictor variables with p values ≤ 0.10 in bivariate analysis were entered into linear regression models. As sensitivity analyses a generalized estimating equations approach was used to account for clustering by center. A p value <0.05 was considered significant in all analyses. Analyses were conducted using SPSS (version 28; SPSS Inc., Chicago, IL) and SAS (v9.4, SAS Institute Inc., Cary, NC).

## RESULTS

### Population

The study population included 890 adults who attended an outpatient CVGC appointment and completed both pre- and post-CVGC study questionnaires. These participants were derived from 3420 patients identified as eligible to participate by the MCVGG-GO centers. Among these, 1322 (40.8%) completed a pre-visit questionnaire. Seventy-one of these patients did not attend their CVGC appointment so were not sent a post-visit questionnaire. The post-CVGC questionnaire was returned by 890/1251 (71.1%) patients who completed the pre-visit questionnaire and attended their CVGC appointment.

**Table 1** summarizes demographic and clinical characteristics of the 890 patients who completed the study. Slightly more than half (497/890, 55.8%) were women and 757 (85.1%) self-reported as White. Clinical indications included cardiomyopathy (436; 46.9%), aortopathy/vasculopathy (187; 21.0%), dyslipidemia (158; 17.8%), and arrhythmias/channelopathies (89; 10%). Most participants (715; 80.3%) were clinically affected. Twenty-one (2.3%) participants were parents attending CVGC for a minor child’s care.

**Table 1.**
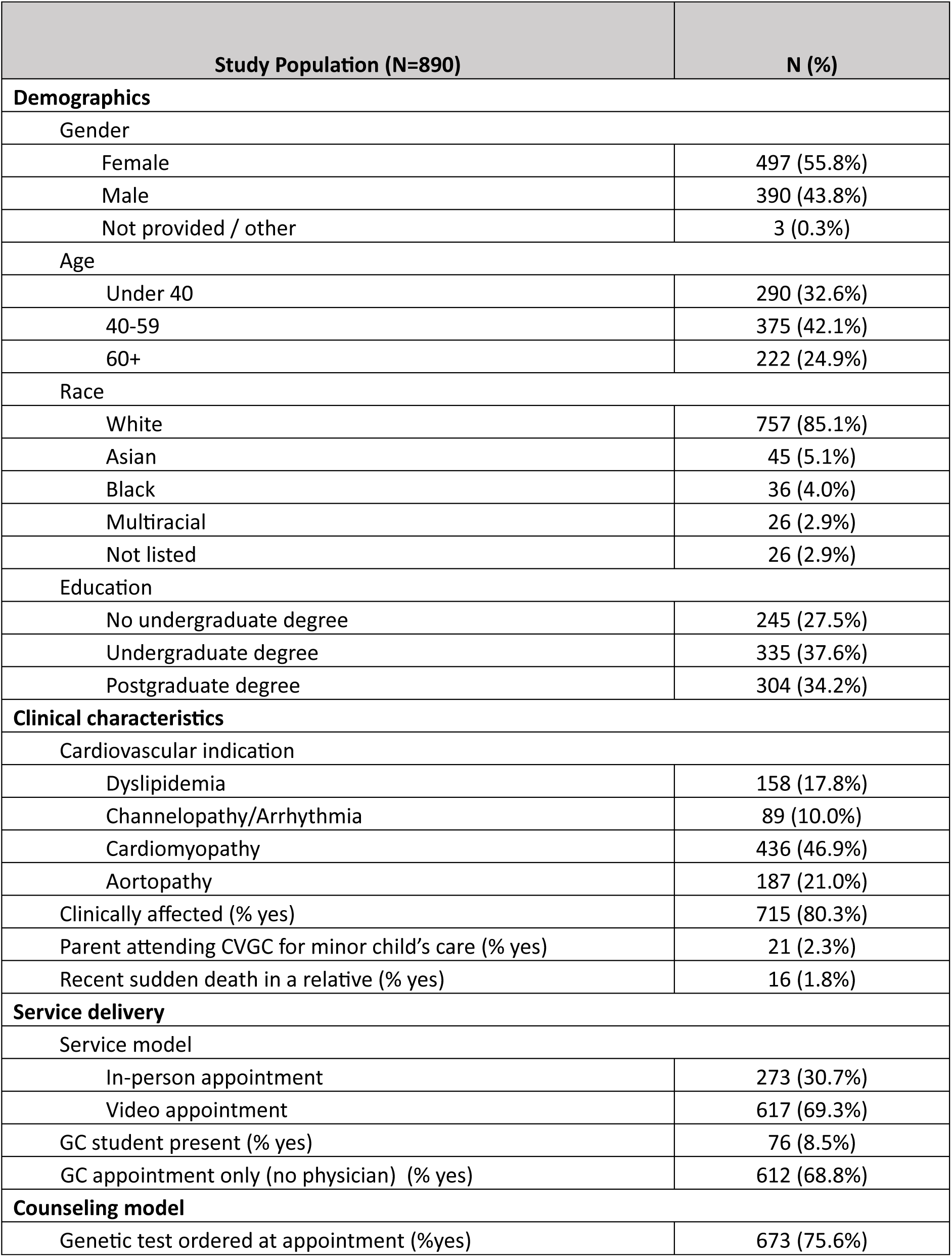
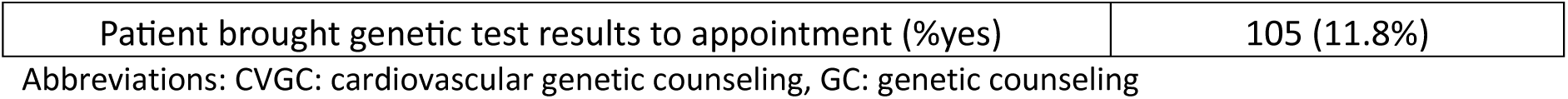
Participant demographic, clinical, and cardiovascular genetic counseling characteristics.

| Study Population (N=890) |  | N (%) |
| --- | --- | --- |
| **Demographics** |  |  |
| Gender |  |  |
| Female |  | 497 (55.8%) |
| Male |  | 390 (43.8%) |
| Not provided / other |  | 3 (0.3%) |
| Age |  |  |
| Under 40 |  | 290 (32.6%) |
| 40-59 |  | 375 (42.1%) |
| 60+ |  | 222 (24.9%) |
| Race |  |  |
| White |  | 757 (85.1%) |
| Asian |  | 45 (5.1%) |
| Black |  | 36 (4.0%) |
| Multiracial |  | 26 (2.9%) |
| Not listed |  | 26 (2.9%) |
| Education |  |  |
| No undergraduate degree |  | 245 (27.5%) |
| Undergraduate degree |  | 335 (37.6%) |
| Postgraduate degree |  | 304 (34.2%) |
| **Clinical characteristics** |  |  |
| Cardiovascular indication |  |  |
| Dyslipidemia |  | 158 (17.8%) |
| Channelopathy/Arrhythmia |  | 89 (10.0%) |
| Cardiomyopathy |  | 436 (46.9%) |
| Aortopathy |  | 187 (21.0%) |
| Clinically affected (% yes) |  | 715 (80.3%) |
| Parent attending CVGC for minor child's care (% yes) |  | 21 (2.3%) |
| Recent sudden death in a relative (% yes) |  | 16 (1.8%) |
| **Service delivery** |  |  |
| Service model |  |  |
| In-person appointment |  | 273 (30.7%) |
| Video appointment |  | 617 (69.3%) |
| GC student present (% yes) |  | 76 (8.5%) |
| GC appointment only (no physician) (% yes) |  | 612 (68.8%) |
| **Counseling model** |  |  |
| Genetic test ordered at appointment (%yes) |  | 673 (75.6%) |

|  |  |
| --- | --- |
| Patient brought genetic test results to appointment (%yes) | 105 (11.8%) |

Abbreviations: CVGC: cardiovascular genetic counseling, GC: genetic counseling

As shown in **Table 1**, the majority of CVGC appointments involved pre-test genetic counseling (673; 75.6%) - e.g., a genetic test was ordered at the visit. Two-thirds (617; 69.3%) of CVGC appointments occurred via video visit. In 76 (8.5%) CVGC appointments a GC student was present and provided at least some genetic counseling.

As shown in **Supplementary Table 1**, participants were seen most often at Johns Hopkins University, Stanford University, and Cleveland Clinic. While these centers accounted for nearly three-quarters of the CVGC visits (652/890; 73.2%), they also employed the majority (15/24; 63%) of the participating GCs.

### Cardiovascular genetic counseling outcomes

**Supplementary Table 2** summarizes GCOS-24, CWS, and CAQ scale and subscale scores pre- and post CVGC and change in each following CVGC. Following CVGC, empowerment (GCOS-24) increased significantly (116.2±13.5 pre-GC to 121.9±13.3 post-GC; +5.94±11.6, p<0.001, Cohen’s *d*: 0.51), cardiac worry (CWS) decreased significantly (15.9±5.01 pre-GC to 15.6±4.67 post GC; +0.40±3.55, p=0.002, Cohen’s *d*: −0.11), and there was no significant change in overall cardiac anxiety (CAQ) (1.43±0.65 pre-GC to 1.44±0.63 post-GC; +0.013±0.34, p=0.29, Cohen’s *d*: 0.04). There were offsetting changes in CAQ subscale scores: participants reported increased cardiac attention (0.078±0.43, p<0.001), decreased cardiac fear (−0.034±0.46, p=0.033), and no significant change in avoidance behaviors (0.030±0.61, p=0.15) post-CVGC. As there was no change in CAQ score observed overall, and the effect sizes of the changes in subscale scores were small, further analyses were limited to the primary study outcomes.

### Association of clinical, demographic, and service delivery characteristics with CVGC outcomes

**Table 2** shows the univariate association of change in GCOS-24 and CWS scale scores with demographic, clinical, and CVGC characteristics. Change in worry (CWS) was associated with patient characteristics and clinical context. The greatest reductions in cardiac worry were observed in patients with the least formal education (p=0.0018). Those without an undergraduate degree had the greatest reduction in worry (−1.09 ± 3.98) while patients with postgraduate degrees had no change in CWS score (0.01 ± 3.19). Age, gender, and race were not associated with change in CWS score. Additionally, patients coming to CVGC with a channelopathy / arrhythmia clinical indication had the greatest reduction in worry (−1.52 ± 4.61) while much more modest reductions were observed for dyslipidemia and cardiomyopathy patients and a slight average increase was seen for patients with an aortopathy indication (p-value for indication=0.007). Additionally, cardiac worry decreased more in at-risk individuals following CVGC than in affected patients (−1.05±3.72 at-risk vs. −0.23 ± 3.47 affected, p=0.010).

**Table 2.** Association of demographic, clinical, and cardiovascular genetic counseling characteristics with extent of change in cardiac worry (CWS scale) and empowerment (GCOS-24 scale).

| Variables | CWS change<br>(Mean ± SD) | p-value | GCOS-24<br>change<br>(Mean ± SD) | p-value |
| --- | --- | --- | --- | --- |
| **Demographics** |  |  |  |  |
| Sex (% female) | -0.56 ± 3.56 | 0.15 | 5.83±11.9 | 0.74 |
| Sex (% male) | -0.20 ± 3.54 | | 6.19±11.1 | |
| Age |  |  |  |  |
| Under 40 | -0.65 ± 3.72 | 0.16 | 5.18±11.0 | 0.29 |
| 40-59 | -0.40 ± 3.51 | | 6.71±12.0 | |
| 60+ | -0.01 ± 3.30 | | 5.13±11.6 | |
| Race |  |  |  |  |
| White | -0.48 ± 3.59 | 0.29 | 6.27±11.4 | 0.10 |
| Asian | -0.50 ± 3.15 | | 5.70±13.1 | |
| Black | 0.58 ± 3.48 | | 1.13±14.3 | |
| Multiracial | 0.68 ± 3.44 | | 2.85±8.16 | |
| Not listed | -0.52 ± 3.19 | | 7.47±12.0 | |
| Education |  |  |  |  |
| No undergraduate degree | -1.09 ± 3.98 | **0.0018** | 5.16±10.9 | 0.43 |
| Undergraduate degree | -0.22 ± 3.42 | | 5.80±11.5 | |
| Postgraduate degree | 0.01 ± 3.19 | | 6.59±12.0 | |
| **Clinical characteristics** |  |  |  |  |
| Indication |  |  |  |  |
| Dyslipidemia | -0.24 ± 3.17 | **0.007** | 6.08±12.1 | 0.74 |
| Channelopathy/Arrhythmia | -1.52 ± 4.61 | | 4.93±11.2 | |
| Cardiomyopathy | -0.42 ± 3.50 | | 5.71±11.8 | |
| Aortopathy | 0.14 ± 3.23 | | 6.69±10.9 | |
| Personal history |  |  |  |  |
| Affected | -0.23 ± 3.47 | **0.01** | 6.14±11.6 | 0.41 |
| At-risk | -1.05±3.72 | | 5.24±11.6 | |
| Sudden death indication |  |  |  |  |
| Yes | -0.33±4.62 | 0.83 | 5.81±11.7 | 0.94 |
| No | -0.071±3.23 | | 6.05±11.6 | |
| **Service delivery** |  |  |  |  |
| GC student present |  |  |  |  |
| Yes | 0.34 ± 3.03 | 0.076 | 3.72±10.7 | 0.092 |
| No | -0.48± 3.58 | | 6.26±11.6 | |
| GC appointment only (no physician) |  |  |  |  |
| Yes | -0.60 ± 3.63 | **0.018** | 5.63±11.4 | 0.28 |
| No | 0.04± 3.35 |  | 6.67±12.0 |  |
| Video or in-person visit |  |  |  |  |
| Video visit | -0.53 ± 3.62 | 0.067 | 5.74±11.3 | 0.47 |
| In-person | -0.03 ±3.27 |  | 6.42±12.1 |  |
| **Counseling model** |  |  |  |  |
| Genetic test ordered at appointment |  |  |  |  |
| Yes | -0.42 ± 3.65 | 0.59 | 5.83±11.3 | 0.70 |
| No | -0.26±3.34 |  | 6.22±12.5 |  |
| Patient brought genetic test results |  |  |  |  |
| Yes | 0.04 ± 3.65 | 0.19 | 7.05±9.85 | 0.29 |
| No | -0.46±3.53 |  | 5.80±11.8 |  |

Abbreviations: GC: genetic counseling

In contrast, change in empowerment was unrelated to clinical indication. Although patients across all race, gender, and age-based subgroups had positive changes in GCOS-24 score (e.g. increased empowerment), there was a trend towards differential extent of increased empowerment by race (p=0.10) with patients identifying as Black having smaller increases in empowerment than patients identifying as White (pairwise comparison (1.13±14.3 Black vs. 6.27±11.4 White; p<0.05 for pairwise comparison with family-wise error rate correction).

Of interest, there was no difference in change in either worry or empowerment associated with genetic test history or ordering. Changes in CWS and GCOS-24 scale scores were similar regardless of whether or not the patient brought in results of a genetic test or whether a genetic test was ordered during the visit.

### Association of the strength of GC:patient alliance with CVGC outcomes

Next, we tested the association of the quality of the GC:patient alliance as rated by the patient via the WAI-SR with study outcomes. **Supplementary Table 2** summarizes WAI-SR whole scale, and goals, task, and bond subscale scores. The distribution of WAI-SR scores was right skewed (towards more favorably rated alliance scores) so divided into quartiles for analysis. Notably, there was no difference between WAI-SR quartile ratings for CVGC visits that included student counselors and those that did not (p=0.98). As shown in **Figure 1**, WAI-SR quartile was positively associated with change in GCOS-24 score following CVGC (GCOS-24 change for 1st quartile WAI-SR score=2.30±12.47, 2nd quartile=6.96±10.15, 3rd quartile=6.83±10.49, 4th quartile=8.59±11.85; p<0.0001). The association of a higher rated working alliance with a larger increase in empowerment was observed for each WAI-SR subscale (**Supplementary Table 3**). The bond subscale was most closely associated with the extent of increase in empowerment (Pearson correlation coefficients: whole scale: 0.222, bond subscale: 0.219; task subscale: 0.181; goal: 0.178; all p<0.0001). In contrast, there was no association between WAI whole scale or any subscale score and extent of CWS score change.

**Figure 1:**
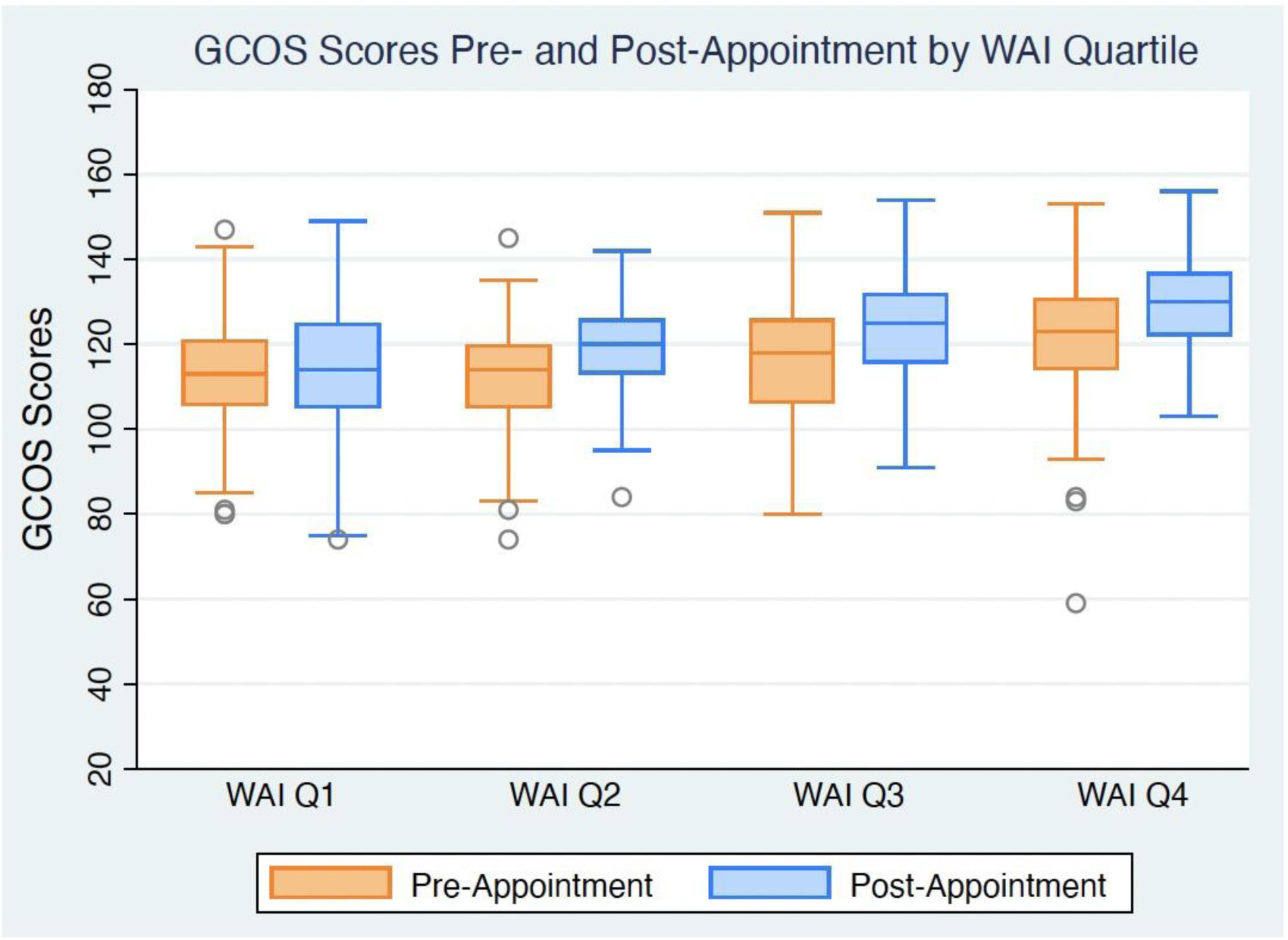
Genetic counseling outcome scale-24 scores pre- and post-cardiovascular genetic counseling **Figure Legend:** Genetic Counseling Outcome Scale-24 (GCOS-24) score prior to (orange) and after (blue) outpatient cardiovascular genetic counseling stratified by patient reported strength of the genetic counselor-patient alliance on the Working Alliance Inventory-SR (modified). Increased empowerment was positively associated with higher ratings of the genetic counselor-patient alliance (p<0.001 – ANOVA). See Supplementary Table 3 for raw values.

### Independent predictors of CVGC outcomes

Multivariable linear regression analyses were performed to identify the independent associations of demographic, clinical, and CVGC characteristics, including the GC:patient alliance, with change in empowerment **(Table 3)** and cardiac worry **(Table 4).**

**Table 3.** Multivariable predictors of change in empowerment (change in GCOS-24 scale score)

| Variable | Estimate | 95% Confidence Interval | P-value | P-value (overall) |
| --- | --- | --- | --- | --- |
| GC student counseling | -3.33 | (-6.66, -0.01) | 0.049 | 0.049 |
| WAI-SR quartile (reference: Quartile 1) |  |  |  | <0.0001 |
| WAI-SR Fourth Quartile | 5.75 | (3.28, 8.22) | <.0001 |  |
| WAI-SR Second Quartile | 4.34 | (1.75, 6.63) | 0.001 |  |
| WAI-SR Third Quartile | 4.16 | (1.64, 6.69) | 0.0012 |  |
| Race (reference: White) |  |  |  | 0.14 |
| Asian | -0.15 | (-3.93, 3.63) | 0.94 |  |
| Black | -4.60 | (-8.67, -0.53) | 0.027 |  |
| Multiracial | -1.35 | (-7.07, 4.40) | 0.65 |  |
| Not listed | 4.04 | (-2.13, 10.2) | 0.20 |  |

Note: Positive estimates reflect greater increases in empowerment post-GC based on GCOS-24 scale score relative to the reference category. Negative values reflect less increase in empowerment relative to the reference category, N=618.
Abbreviations: GC: genetic counseling, WAI-SR: Working Alliance Inventory – Patient version

**Table 4:**
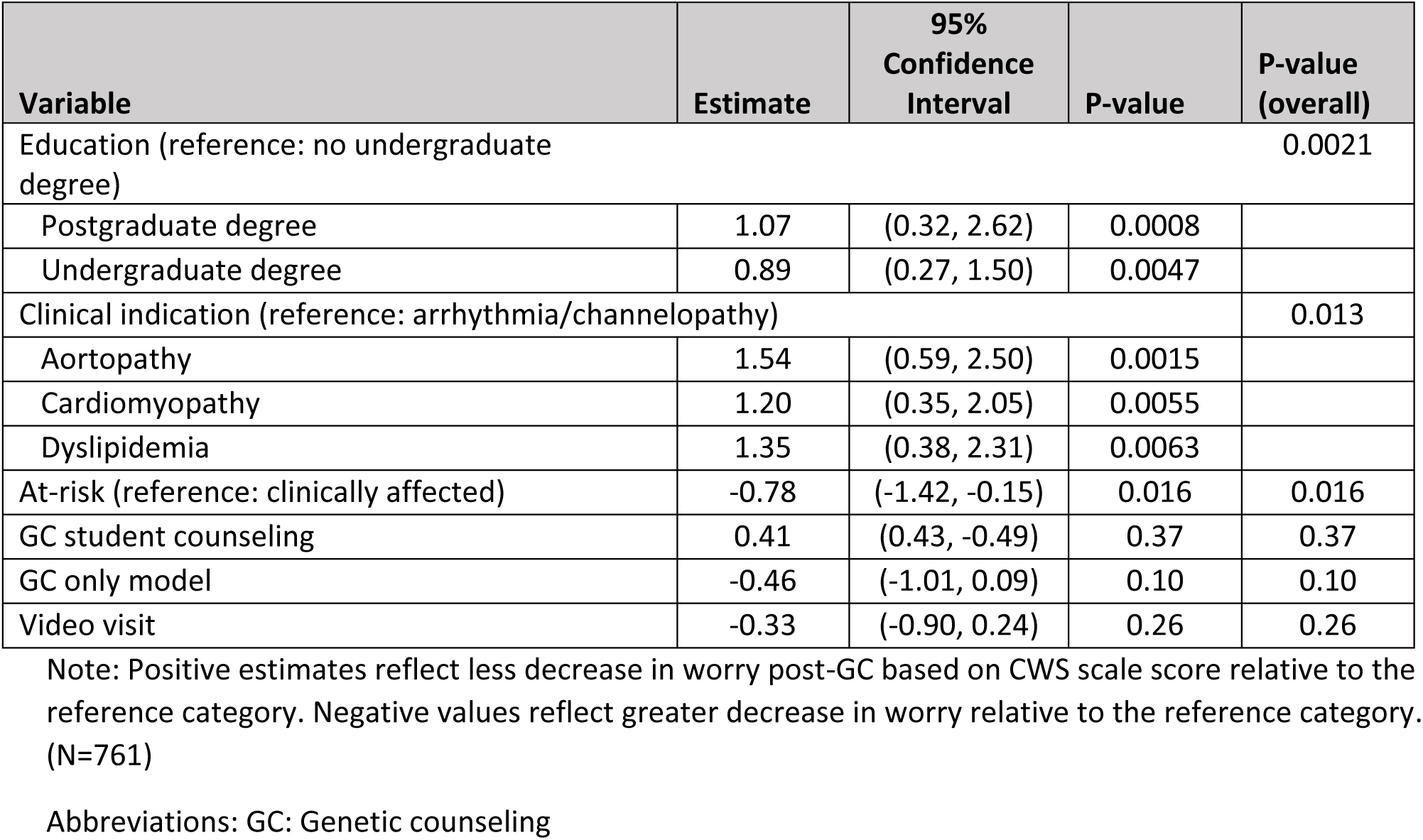
Predictors of change in worry (CWS scale score)

In multivariable analysis, strength of the GC:patient alliance, student genetic counseling, and race were independently associated with change in empowerment. WAI-SR ratings in the lower quartiles were associated with significantly smaller increases in empowerment (p<0.0001; type 3 analysis). Student counseling was independently associated with smaller increases in empowerment than CVGC performed solely by board certified GCs (p=0.049).

Finally, patients self-identifying as Black had smaller gains in empowerment than those identifying as White (p=0.027), although race overall was not independently associated with change in empowerment (p=0.14, type 3 analysis). In a sensitivity analysis controlling for enrolling center, similar findings were observed although statistical significance was attenuated (**Supplementary Table 4**).

In multivariable regression, patient education and clinical indication were associated with CWS change. At-risk individuals had significantly greater reductions in worry than clinically affected individuals (p=0.016). Additionally, patients with an arrhythmia indication or (suspected) channelopathy diagnosis had significantly greater reductions in worry following

CVGC than patients with an indication associated with structural heart disease (p=0.013; type 3 analysis). Finally, patients with less formal education experienced significantly greater reductions in worry (p=0.002; type 3 analysis). As with change in GCOS-24, results accounting for clustering within center showed attenuation of statistical significance (**Supplementary Table 5**).

## DISCUSSION

In this large multicenter, multi-provider, observational study we found that CVGC was associated with decreased worry and increased empowerment prior to the return of any genetic test results ordered in conjunction with the GC session. Thus, this study provides foundational evidence that CVGC is associated with improved psychosocial patient metrics separate from the impact of genetic test results. Furthermore, as the combination of increased empowerment and decreased worry is expected to support improved health outcomes, these results provide evidence to support guideline-recommended inclusion of CVGC in the care of families with IHD^1,13,37^. These findings also provide a nascent evidence base to inform design and evaluation of future CVGC service delivery.

These results confirm and extend findings from prior CVGC outcomes studies. We found that the strength of the relationship between the GC and the patient is strongly associated with the extent of empowerment change after CVGC across cardiovascular clinical indications, consistent with a prior study of CVGC for arrhythmogenic cardiomyopathy ^13^. As empowerment is one of the goals of genetic counseling and is associated with improved psychosocial and potentially health-related adherence outcomes, this is of particular importance in guiding CVGC practice. Additionally, the strength of the provider and patient relationship, or alliance, which tracks with improvements in empowerment, has been associated with improving symptoms and complaints, and positive health behaviors including treatment adherence^20^.

We showed that worry decreased after CVGC, especially in those in with less formal education. There is an extensive body of literature regarding psychosocial distress including anxiety, depression, and lower quality of life in IHD families^5^. This reduction in worry is similar to the impact of genetic counseling reported in oncology populations^25^. Additionally, we found that the extent of reduction in worry varied by clinical indication and was especially large in the arrhythmia referral subgroup and in those at risk vs. those with an already established clinical diagnosis. This variability is not entirely unexpected, as in some cases, CVCG does inform patients of risks and appropriate management strategies, which can increase worry in some populations. Our findings were also are in line with previous work showing a modest reduction in the CAQ fear subscale (−0.034±0.46)^13^. Put together, these findings highlight the potential of CVGC to reduce worry and fear in those at risk for sudden cardiac death and with less formal education. The lack of association of reduction in worry with the GC:patient alliance suggests that alternative approaches to CVGC service delivery other than 1:1 genetic counseling by a GC may be similarly effective at ameliorating worry^38^.

Across clinical genetics practice, there is a struggle to differentiate clinical utility and positive patient health outcomes related to the genetic test implications and genetic counseling. Separating these two interventions is critical in evaluating the value of CVGC. We found that CVGC is associated with patient benefit prior to the return of any genetic test results ordered at the visit. This study is also one of few to specifically assess the differences in outcomes related to timing of the genetic testing relative to CVGC. There was no difference in change in either empowerment or worry based on the timing of CVGC appointment relative to genetic testing. Thus, not only were increased empowerment and decreased worry attributable to the CVGC visit itself rather than the genetic test, but in the era of more commonplace ordering of genetic testing outside of specialty clinics (e.g. mainstreaming), the post-test CVGC visit is expected to continue to deliver important patient benefit.

### Clinical implications

This clinically vital work has replicated and extended previous work suggesting that CVGC provided by board certified/eligible cardiovascular GCs, is associated with improved psychosocial outcomes, specifically increases in empowerment, and reductions in worry and fear. Empowerment, especially strengthened by the GC:patient alliance, is of particular importance as trust in medical recommendations and adherence is critical in preventing sudden death or other poor outcomes of IHD (e.g., heart failure). High cardiac anxiety has been associated with worse functional and physical outcomes, and therefore reduction in anxiety or worry in this population can be linked to better physical and psychological outcomes^29^. All of this contributes to the evidence base supporting the value of CVGC and supports guideline recommendations in encouraging referral practices and payor reimbursement.

### Implications for future research

These data also have the potential to inform future CVGC research. Our results demonstrate the performance of clinically relevant patient-reported outcome measures in a CVGC context and provide data that can be used to estimate expected effect sizes for common CVGC clinical indications. Furthermore, these results suggest which outcomes are sensitive to the quality of the CVGC relationship (e.g., empowerment) and which may be more amenable to alternative service delivery. Such insights can be used to inform design of interventions and trials that test them.

### Limitations

This study is both strengthened and limited by the fact that data were derived from existing CVGC clinics/practice and characterize usual care. As an observational study, there was no standard genetic counseling script or template for the visits. Furthermore, visits were not recorded. There may be variability in the content and psychotherapeutic approach to CVGC among centers and individual GCs that could affect outcomes that were thus not measured. In addition, small numbers with characteristics of interest within centers (e.g., multiple circumstances of zero cases in a center) led to reduced power in analysis accounting for clustering by center. There is also no long-term follow up data to assess the longevity of these outcomes or to explore the impact of CVGC on how genetic test results are perceived. A modest response rate furthermore suggests there were patient experiences that were not well-represented in the dataset.

### Conclusions

This is the first large, multicenter assessment of CVGC outcomes across clinics, indications, and countries. A CVGC visit is associated with increased patient empowerment, particularly in the context of a strong relationship between the GC and the patient, independent of results of any genetic testing ordered. CVGC is also associated with less worry and fear, especially in patients with less formal education, and in those undergoing cascade genetic testing for risk of IHD. These data thus suggest that the process inherent in CVGC has positive health outcomes outside of the outcomes related to the genetic test. Future work is needed in more diverse ancestries and populations, and to confirm these positive outcomes in emerging more efficient CVGC models.

## Data Availability

Because of the sensitive nature of the data collected and due to restrictions on data sharing the full dataset will not be made available. Requests to access limited data elements from qualified researchers trained in human subject confidentiality protocols may be sent to the corresponding author.

## Acknowledgments

The authors would like to thank the inherited heart disease families who participated in this work. CJ and BM would like to acknowledge Shannon McKee who aided in figure and table development. CR and HI would like to thank Daniela Diaz Caro, Beatriz Anguiano, Jake Plewa and Kendall Schmidt who aided in data collection. EW and SW would like to acknowledge Stephanie Harris who aided in recruitment and Lulwa El-Saket who assisted in data collection. LY is the recipient of a co-funded National Heart Foundation of Australia/ National Health and Medical Research Council (NHMRC) PhD Scholarship (#102568/#191351). JI is the recipient of a National Heart Foundation of Australia Future Leader Fellowship (#106732).

